# Assessing Body Composition via a Smartphone Computer Vision Application: Reliable but Biased Compared to BODPOD and InBody

**DOI:** 10.1101/2025.08.01.25332763

**Authors:** Jahliya Sisohor, Jatin Ambegaonkar, Bryndan Lindsey, Yosef Shaul, Joel Martin

## Abstract

Accurate assessment of body composition is essential for monitoring health status, fitness progress, and disease risk. Traditional methods such as air displacement plethysmography (BODPOD) and bioelectrical impedance analysis (BIA) are widely used to estimate body fat percentage (BF%), fat mass (FM), and fat-free mass (FFM), but can be costly or inaccessible. Smartphone applications utilizing computer vision (CV) offer a promising alternative. This study evaluated the agreement and test-retest reliability of a smartphone-based CV application (CV_app_) compared to BODPOD and the InBody BIA device. Forty-nine adults (ages 18–70; 29 females, 27 racial and ethnic minority participants) completed two consecutive measurements using BODPOD, InBody, and CV_app_ in a single session. Differences in BF%, FM, and FFM estimates were analyzed using repeated-measures ANOVA. Agreement metrics included mean absolute error (MAE), root mean square error (RMSE), concordance correlation coefficients (CCC), and Bland-Altman analysis. Subgroup analyses examined differences by sex and minority status. The CV_app_ yielded significantly higher BF% (mean=+2.2%, *p*=0.004) and FM (+1.5 kg, *p*=0.015) compared to BODPOD, and lower FFM than InBody (mean=−1.84 kg, *p*=0.043). The CV_app_ agreement with BODPOD (MAE=4.0, RMSE=5.0, CCC=0.86) was lower than with the InBody (MAE=3.3, RMSE=4.3, CCC=0.89), with wider limits of agreement. All methods showed excellent test-retest reliability (ICC>0.99). A significant Sex×Method interaction was observed for BF% (*p*<0.001), FM (*p*<0.001), and FFM (*p*=0.013), with females showing greater overestimation of BF% (mean=+3.9%) and FM (mean=+2.7kg, *p*<0.001), and underestimation of FFM (mean=−2.5kg, *p*<0.001) by the CV_app_. Among minority participants, BF% and FM estimates from the CV_app_ were significantly higher than BODPOD (mean=+2.7%, *p*=0.009; mean=+1.8kg, *p*=0.031), and FFM was lower (mean=−1.8kg, *p*=0.035), with significant Method×Minority Group interactions for FM and FFM (*p*<0.03). While the CV_app_ was reliable, its estimates differed from other common body composition methods and revealed population-specific biases, highlighting the need for more accurate and equitable algorithms.

## Introduction

Body composition, which is the proportion of fat (BF%), muscle, and other tissues in the body, is a key indicator of health [1] and physical performance [2,3]. Excess adiposity (*e.g*., obesity) is a well-recognized risk factor for chronic diseases such as type 2 diabetes, cardiovascular disease, and certain cancers [1]. The prevalence of obesity has risen dramatically and as of 2022, an estimated 16% of adults worldwide (∼890 million people) were living with obesity [1]. In the United States, obesity now affects over 2 in 5 adults (41.9% in 2017–2020), reflecting a significant increase from previous decades [4]. Conversely, low BF% or low muscle mass can also carry health implications (*e.g.* malnutrition or sarcopenia) [2], further emphasizing that body composition is central to health status. Beyond health and disease, body composition is crucial for performance in athletic [5,6] and occupational settings [3,7,8]. In clinical practice and research, having reliable methods to quantify body composition allows practitioners to better evaluate nutritional status [9], tailor interventions for weight management [10], or strength training [11], and predict functional outcomes [2,5].

A variety of methods are available to assess body composition, each with strengths and limitations [12], Historically, underwater weighing (*e.g.,* hydrostatic densitometry) was regarded as a gold-standard two-compartment model for body composition measurement [9,12]. However, due to practical limitations of hydrostatic weighing several methods have become more frequently used. One method is air displacement plethysmography (ADP), performed most commonly with the commercial BODPOD system. The BODPOD measures body volume by air displacement while a participant sits in a sealed chamber, and it computes body density and BF% using known equations (*e.g.* Siri equation) [13]. The method is quick and non-invasive, and its accuracy in determining body composition is generally considered high [13]. For instance, ADP BF% estimates typically agree within a few percentage points of more direct methods [13]. Nonetheless, ADP has known sources of error [13]. Factors like clothing, air trapped in hair, or moisture on skin can affect the volume measurement and thus impact accuracy [13].

Another widely used body composition assessment method is bioelectrical impedance analysis (BIA) [14]. BIA estimates body composition by sending a low electrical current through the body and measuring the impedance, which varies between lean tissue (*i.e.,* high water content, more conductive) and adipose tissue (*i.e.,* low water, more resistive). Advantages of BIA include its portability, speed, and relatively low cost, making it one of the most popular methods in both clinical and fitness settings [15]. However, accuracy is a major concern with BIA [16]. The consistency of BIA results compared to reference techniques is variable as studies report that BIA can significantly diverge from ADP or dual-energy X-ray absorptiometry (DXA) measurements [14,17]. For example, BF% measured by BIA using the InBody 770 has been shown to be underestimated, on average, compared to values obtained from the BODPOD in young adults [16]. Moreover, BIA accuracy can be influenced by a person’s hydration status, recent food or exercise, and even the specific device algorithms used [14]. In addition to ADP and BIA, other techniques, such as DXA, Magnetic Resonance Imaging, or Computerized Tomography scans, can provide more detailed body composition data, including regional fat distribution and muscle mass, but these are costly, not portable, and typically limited to clinical or research facilities [12]. Thus, there is a need for methods that are both accurate and accessible.

Given the limitations of traditional techniques, recent efforts have turned to computer vision and digital anthropometry that leverage smartphone cameras to estimate body composition from digital images [18,19]. The theoretical foundation of computer-vision based approaches lies in the relationship between external body shape and internal composition [20]. Modern computer vision algorithms can extract anthropometric information, such as body dimensions, contours, and volumes, from photographs. For example, computer vision algorithms can construct a 3D (3-Dimensional) avatar of a person from a series of smartphone digital photos, then calculate circumferences, volumes, and other shape metrics to estimate body composition metrics [21].

The benefits of computer vision-based body composition tools are substantial as they require minimal equipment – typically just a smartphone camera without specialized hardware. Such tools are also quick and non-invasive. For instance, a user can obtain results in minutes with only a few photos, avoiding the discomfort of BODPOD enclosure or the exposure to X-rays from DXA. Most computer vision approaches to estimate body composition use machine learning to predict body fat or muscle metrics from image features [19]. For example, convolutional neural networks (CNNs) can be trained on thousands of labeled images to recognize subtle visual patterns of adiposity [22]. Other methods fit a statistical body shape model to the image, which can then be used to estimate tissue volumes [20,23].

Several studies have evaluated the accuracy and reliability of smartphone or camera-based body composition estimation. Farina et al. [24] developed a smartphone computer vision application (CV_app_) that analyzes a single lateral full-body photograph to estimate fat mass (FM). In a sample of 117 adults, the application’s FM estimates showed a strong association with DXA (*R*^2^=0.95 for females, 0.91 for men) and no significant bias [24]. Affuso et al. [23] reported a method using two digital images per person combined with a support vector regression machine learning algorithm to predict BF%. In 226 adults, the photo-based model achieved a correlation of *r*=0.87 with DXA BF%. The mean BF% by photography was within 0.1% of the DXA value in adults, indicating excellent agreement [23]. Tian et al. [20] introduced an algorithm to predict 3D body shape and composition from a single 2D photograph. The method used a frontal image to extract a silhouette, fit a 3D statistical shape model, and then estimated body composition from the 3D avatar. When evaluated against DXA in a validation set, the system showed no significant difference between the photo-predicted and DXA-measured values of total body fat, and achieved high correlations (*R*^2^=0.81 in men, 0.74 in female for BF%). Importantly, Graybeal and colleagues found that for 3D scanning devices the validity varied between sexes and racial and ethnic groups [18]. This provides some evidence that demographic factors could influence body composition measurement accuracy when using digital imaging approaches.

More recently, studies have directly compared smartphone-based body composition tools to gold standards and other field methods [19,25,26]. Majmudar et al. [26] validated a smartphone CVapp using CNNs in 134 adults, comparing it to DXA and multiple BIA devices, including smart scales and the BODPOD. The app showed high concordance with DXA (CCC = 0.93–0.96), no significant bias, and the narrowest limits of agreement (±5% fat), outperforming other methods that showed greater bias and variability. In addition to accuracy, the reliability of smartphone-based body composition measures has been examined. Metoyer et al. [25] conducted an agreement study to test a smartphone body composition CV_app_ under various real-world conditions, such as different smartphone camera resolutions, lighting levels, and background colors. Interestingly, there was an effect of background color on BF% estimates, with non-white backgrounds yielding higher BF% estimates as compared to the white background condition. However, the BF% estimates under all tested conditions were strongly correlated with those under the reference condition [25].

Collectively, the literature suggests that smartphone-based CV_app_ methods for body composition can be accurate and reliable [19,25–27]. However, most prior studies have validated these tools against DXA [20,24,26] with fewer data existing on how a CV_app_ compares with other common methods like BODPOD and BIA or on the test–retest reliability of a CV_app_. Importantly, few studies have examined whether the accuracy and reliability of smartphone-based CV_app_ assessments are consistent across sexes and among individuals from racial and ethnic minority groups, an important consideration for equitable adoption. Therefore, the purpose of the current study is to assess the agreement and reliability of a smartphone-based body composition CV_app_ in comparison to two common methods, ADP (*i.e.,* BODPOD) and BIA (*i.e.,* InBody), used in research, clinical and other settings. Specifically, this study aims to: 1) Compare body composition (BF%, FM, fat-free mass [FFM]) estimates from the CV_app_ to those from BODPOD and BIA, evaluating the level of agreement and identifying any systematic differences between methods; 2) Determine the test–retest reliability of the CV_app_, by examining the consistency of BF% measurements across consecutive tests under standardized conditions; and 3) Examine potential differences in accuracy and reliability of the CV_app_ across sex and racial and ethnic groups to expand upon existing validation efforts. The findings can help inform the responsible and inclusive use of such technology in health, performance, and telemedicine settings.

## Materials and Methods

### 2.1 Study Design

A repeated-measures, single-session design was used to compare body composition estimates across three methods: 1) ADP via the BODPOD, 2) BIA via the InBody, 3) and a smartphone CV_app_. Each participant completed two consecutive measurements per method under standardized laboratory conditions to assess agreement and test-retest reliability. All assessments were conducted in the Sports Medicine Assessment Research & Testing (SMART) Laboratory at XXX (BLINDED FOR REVIEW) University (IRB #: 1665548). The same researcher administered all assessments to minimize interrater variability.

### 2.2 Participants

Forty-nine healthy adults (29 females, 20 males) between the ages of 21 and 70 years (mean ± SD: 37.2 ± 14.6 years) completed the study procedures. Participants’ self-reported race and ethnicity using U.S. Census classifications. A majority of the participants were White (American Indian or Alaskan Native, *n*=0; Biracial/other, *n*=2; Asian, *n*=7; Black or African American, *n*=7; Hispanic or Latino, *n*=8; Middle Eastern or North African, *n*=2; Native Hawaiian or Pacific Islander, *n*=0; White, *n*=22) and for statistical purposes of having sub-groups large enough to perform group comparisons participants were dichotomized as either non-racial and ethnic minorities (*i.e.,* White) or racial and ethnic minorities (*i.e.,* all self reporting as non-White) [28]. Inclusion criteria included age ≥18 years, no current pregnancy, and no implanted pacemakers. Participants were instructed to avoid exercise, alcohol, and excessive caffeine for 24 hours and to fast for 4 hours before testing, except for water, which could be consumed up to 45 minutes prior. The study protocol was approved by the university’s Institutional Review Board and all participants provided written informed consent prior to participation in the study.

### 2.3 Protocol

Upon arrival, participants changed into the required testing attire consisting of black spandex shorts and swim cap with sports bras for females. Each participant’s height and weight were measured using a stadiometer (Detecto, Webb City, MO, USA) and a digital scale (EatSmart, Tokyo, Japan) then these values were recorded and later entered into the devices or app as needed. The body composition assessments were administered in a fixed sequence for all participants with two consecutive measurements of: (1) BODPOD, (2) InBody, and (3) CV_app_ for consistency of methodology and practicality (see Fig 1). For each method, the two measurements were completed consecutively with recalibration of the instrument between trials as appropriate. All CV_app_ measurements were performed in the same laboratory space with a plain white backdrop for photography and consistent ambient lighting. Throughout the session, the researcher followed identical procedures for every participant to minimize any variability in test administration and the same researcher tested all participants.

**Fig 1.**
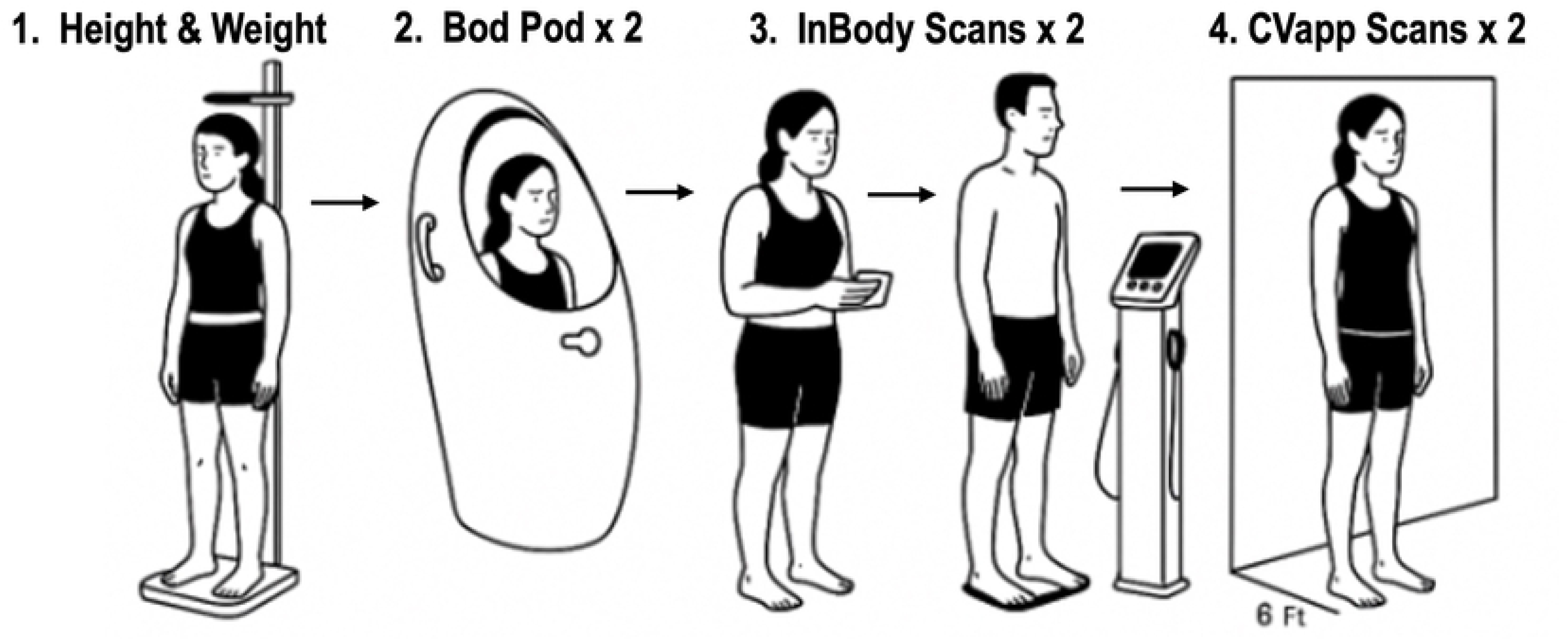
Study protocol (n=49 participants) **Notes:** The body composition assessments were administered in a fixed sequence for all participants, with two consecutive measurements each for the BODPOD, InBody, and computer vision smartphone application. A total of 29 females and 20 males completed all procedures, including 27 participants who self-identified as members of racial and ethnic minority groups.

### 2.4 Body Composition Measurements

#### Air Displacement Plethysmography (ADP)

Body composition was measured using a BODPOD GS-X system (COSMED USA Inc., Concord, CA, USA). The device was calibrated at the start of each day of testing. During each BODPOD trial, the participant sat inside the chamber wearing minimal attire and a swim cap. The system measured body volume twice and averaged the values, with thoracic gas volume predicted by the software. Body density was calculated from body mass and volume, and BF% was then derived using the Siri equation based on the measured body density. Each participant underwent two sequential BODPOD assessment measurements as described above.

#### Bioelectrical Impedance Analysis (BIA)

Body composition was assessed using the InBody 570 device (InBody USA, Cerritos, CA, USA), which employs a tetrapolar 8-point tactile electrode system and multifrequency impedance analysis. Participants stood barefoot on the scale platform and gently held the hand electrodes. Prior to measurement, participants wiped hands and feet with the manufacturer’s electrolytic tissues to improve conduction. The device automatically measured the participant’s weight, while height from the stadiometer measurement was manually input by the researcher. The InBody then analyzed impedance to estimate BF%, FM, and FFM. Two sequential InBody measurements were completed for each participant, with the participant stepping off the platform and then re-resuming the testing position for the second measurement.

#### Computer Vision Smart Phone App

The CV_app_ Body Composition Scanner (Spren Technologies, Asheville, NC, USA) was used on an iPhone 13 Pro Max (Apple Inc., Cupertino, CA, USA) to estimate body composition via digital images obtain with a 12 megapixel (MP) camera. A profile was created for each participant in the app, and demographic data (age, sex, height, weight, and ethnicity) were entered before scanning. The smartphone was fixed on a tripod at a marked distance (1.83 m / 6 feet) and height (0.61 m / 2 feet) from the participant. For each scan, two digital images of the front and right side view were captured with the participant standing upright against a plain white background under natural light. The camera distance was standardized, background, and lighting for all app measurements, as consistent conditions are known to improve the accuracy of image-based body composition analysis (e.g., white background yields higher agreement than darker backgrounds) [25]. The CV_app_’s computer vision algorithm processed the images and generated estimates of BF%, FM, and FFM. Each participant completed two sequential measurements with the CV_app_.

### 2.5 Statistical Analysis

Our analyses focused on assessing agreement between methods, test-retest reliability, and subgroup performance by sex and racial and ethnicity. The three measurement methods (BODPOD, InBody, and CV_app_) were the primary factor of interest while BF%, FM, and FFM served as the outcome variables. Raw values were first screened for missing or implausible entries. Normality was assessed with Shapiro-Wilk tests and visual inspection of data distributions. For each method, test-retest data were averaged for descriptive and agreement analyses. As described above participants were categorized dichotomously into Minority and Non-Minorities based on self-reported race and ethnicity. Descriptive statistics were calculated for participant characteristics and for each body composition outcome by method and demographic variables.

To determine if the three methods yielded significantly different body composition measures, a one-way repeated-measures ANOVA on each outcome (BF%, FM, FFM), with method (3 levels: BODPOD, InBODY, CV_app_) as the within-subjects factor was conducted. Mauchly’s test was used to verify the sphericity assumption for each ANOVA; if violated, a Greenhouse-Geisser correction was used. When a significant main effect of method was found, pairwise post-hoc comparisons between methods were performed with Bonferroni adjustments. Effect sizes for main effects were reported as generalized eta-squared (*η*²_G_) values and interpreted as 0.01 for *small*, 0.06 for *medium*, and 0.14 or greater for *large* effects.[29] Pair-wise effect sizes were reported as Cohen’s d and interpreted as small (*d* = 0.2), medium (*d* = 0.5), and large (*d* = 0.8) [29].

Agreement between methods was evaluated via Bland–Altman analysis: for each pair of methods, we calculated the mean bias as the mean difference in BF%, FM, or FFM between the two methods and the 95% LOA (computed as bias ± 1.96 SD of the differences) to assess the extent of agreement and to check for any systematic bias or proportional bias between methods. In addition, method agreement was further examined using Pearson correlation coefficients (*r*) to assess linear associations, mean absolute error (MAE) and root mean square error (RMSE) to evaluate average and squared deviations, Lin’s CCC to assess accuracy and precision,[30] and intraclass correlation coefficients (ICC) using a two-way model for absolute agreement. CCC values were interpreted as <0.90 = *poor*, 0.90–0.95 = *moderate*, and >0.95 = *excellent* agreement.[31] ICC values were interpreted using the following thresholds: <0.50 = *poor*, 0.50–0.74 = *moderate*, 0.75–0.89 = *good*, and ≥0.90 = *excellent* reliability [32].

Test–retest reliability for each measurement technique was assessed using multiple metrics. For each device/app, the two successive measurements of BF%, FM, and FFM were compared. We computed the ICC (2,1) for each method using a two-way random-effects model (single measures, absolute agreement) to quantify relative consistency between measurement 1 and 2. The standard error of measurement (SEM) and the coefficient of variation (CoV) for each method’s repeated measurements was computed to gauge absolute measurement error and within-method variability. SEM was calculated using the standard deviation (SD) of the differences between two repeated measurements, divided by the square root of two. This approach assumes equal error variance across measurements and provides an estimate of the typical measurement error inherent to the testing procedure. The CoV was calculated to express the SEM as a percentage of the overall mean of both measurements: CoV = (SEM / overall mean) x 100%. The ICC (2,1) was used to evaluate consistency between measurements. The Coefficient of Reliability (CR) was calculated as 1.96 times the SD of the differences, representing the range within which 95% of the differences between measurements are expected to occur.

Subgroup analyses were conducted to examine whether body composition estimates differed by sex (male vs. female) and self-reported racial and ethnic minority status (Minority vs. Non-Minority). A series of repeated-measures ANOVA models were used to test for main effects of method and subgroup, as well as Method × Sex and Method × Minority Group interactions. Greenhouse-Geisser corrections were applied when Mauchly’s test indicated violations of sphericity. Follow-up pairwise comparisons of estimated marginal means were conducted using Tukey’s adjustment. To evaluate potential differences in participant demographics, a chi-square test of independence was used to assess the association between sex and Minority Group status. Statistical significance was set at α < 0.05 for all tests. All statistical analyses were performed using R (version 4.3.1).

## Results

Descriptive statistics of the participants are provided in Table 1. The BODPOD assessment indicated that the average BF% among participants was 24.5% (SD = 9.6%), with values ranging from 3.8% to 47.4%. Males were taller (*t*(48) = -4.89, *p* < 0.001, *d* = 1.40) and had greater body mass than females (*t*(48) = -4.52, *p* < 0.001, *d* = 1.29). In terms of body composition, females exhibited higher BF% (*t*(48) = 3.16, *p* = 0.003, *d* = 0.91) and lower FFM (*t*(48) = -7.94, *p* < 0.001, *d* = 2.28) compared to males. However, FM did not differ between sexes (*t*(48) = 0.77, *p* = 0.45, *d* = 0.22). Participants identifying as members of a Minority Group were younger compared to non-minority participants (*t*(48) = -3.22, *p* = 0.002, *d* = 0.92). No statistically significant differences were observed between Minority and Non-Minority participants for height (*t*(48) = 0.15, *p* = 0.88, *d* = 0.04), body mass (*t*(48) = 0.33, *p* = 0.74, *d* = 0.10), BF% (*t*(48) = -0.42, *p* = 0.68, *d* = 0.12), FM (*t*(48) = -0.26, *p* = 0.80, *d* = 0.07), or FFM (*t*(48) = 0.45, *p* = 0.66, *d* = 0.13).

**Table 1.** Participant Characteristics.

| | All<br>($n=49$)<br>Mean ± SD<br>[Range Min,<br>Range Max] | Males<br>($n=20$) | Females<br>($n=29$) | Racial and<br>Ethnic Non-Minorities<br>($n= 22$) | Racial and<br>Ethnic<br>Minorities<br>($n = 27$) |
| --- | --- | --- | --- | --- | --- |
| Age (years) | 37.2 ± 14.6<br>[21 , 70] | 41.9 ± 15.8<br>[22 , 70] | 33.9 ± 13.1<br>[21 , 68] | 43.8 ± 15.2<br>[22, 70] | 31.8 ± 11.9<br>[22, 70] |
| Height (cm) | 171.3 ± 8.5<br>[158.2, 191.2] | 177.0 ± 7.7<br>[161.5, 191.2] | 167.3 ± 6.7<br>[158.2, 180.5] | 171.3 ± 7.8<br>[158.2, 188.3] | 171.2 ± 9.2<br>[158.9, 191.2] |
| Mass (kg) | 77.2 ± 15.4 | 86.9 ± 13.4 | 70.5 ± 13.1 | 76.9 ± 11.2 | 77.4 ± 18.3 |
|  | [50.6, 123.0] | [71.2, 123.0] | [50.6, 99.5] | [55.5, 99.5] | [50.6, 123.0] |
| BF (%) | 24.6 ± 9.6<br>[3.8, 47.4] | 19.9 ± 8.4<br>[3.8, 38.8] | 27.8 ± 9.2<br>[10.8, 47.4] | 25.1 ± 11.0<br>[8.4, 47.4] | 24.2 ± 8.6<br>[3.8, 38.8] |
| FM (kg) | 19.2 ± 9.8<br>[2.9, 47.6] | 17.9 ± 10.3<br>[2.9, 47.6] | 20.1 ± 9.5<br>[6.0, 47.0] | 19.6 ± 10.3<br>[6.0, 47.0] | 18.9 ± 9.5<br>[2.9, 47.6] |
| FFM (kg) | 57.7 ± 12.3<br>[38.3, 85.9] | 68.7 ± 7.8<br>[55.5, 85.9] | 50.2 ± 8.5<br>[38.3, 73.0] | 57.1 ± 10.3<br>[42.4, 77.3] | 58.2 ± 13.9<br>[38.3, 85.9] |

**Note:** Values are Mean ± standard deviation and minimum and maximum values below in brackets for air displacement plethysmography (e.g., BODPOD). **Abbreviations:** BF%, body fat percentage; FM, fat mass; FFM, fat-free mass; SD, standard deviation; Min, minimum; Max, maximum.

### Agreement Between Methods

A repeated measures ANOVA revealed significant main effects of measurement method on all body composition outcomes (See Table 2). Compared to the BODPOD, the CV_app_ yielded significantly higher estimates of BF% (*p* = 0.005, *d* = 0.47) and FM (*p* = 0.021, *d* = 0.40), and significantly lower FFM (*p* = 0.042, *d* = 0.37). When compared to InBody, the CV_app_ also produced higher estimates of BF% and FM, though these differences were not statistically significant after Bonferroni correction (BF%: *p* = 0.051; FM: *p* = 0.078). The FFM estimate from CV_app_ was significantly lower than that of InBody (*p* = 0.052, *d* = −0.35). In contrast, comparisons between BODPOD and InBody showed no significant differences for BF%, FM, or FFM. A summary of post hoc comparisons is provided in Table 2.

**Table 2.**
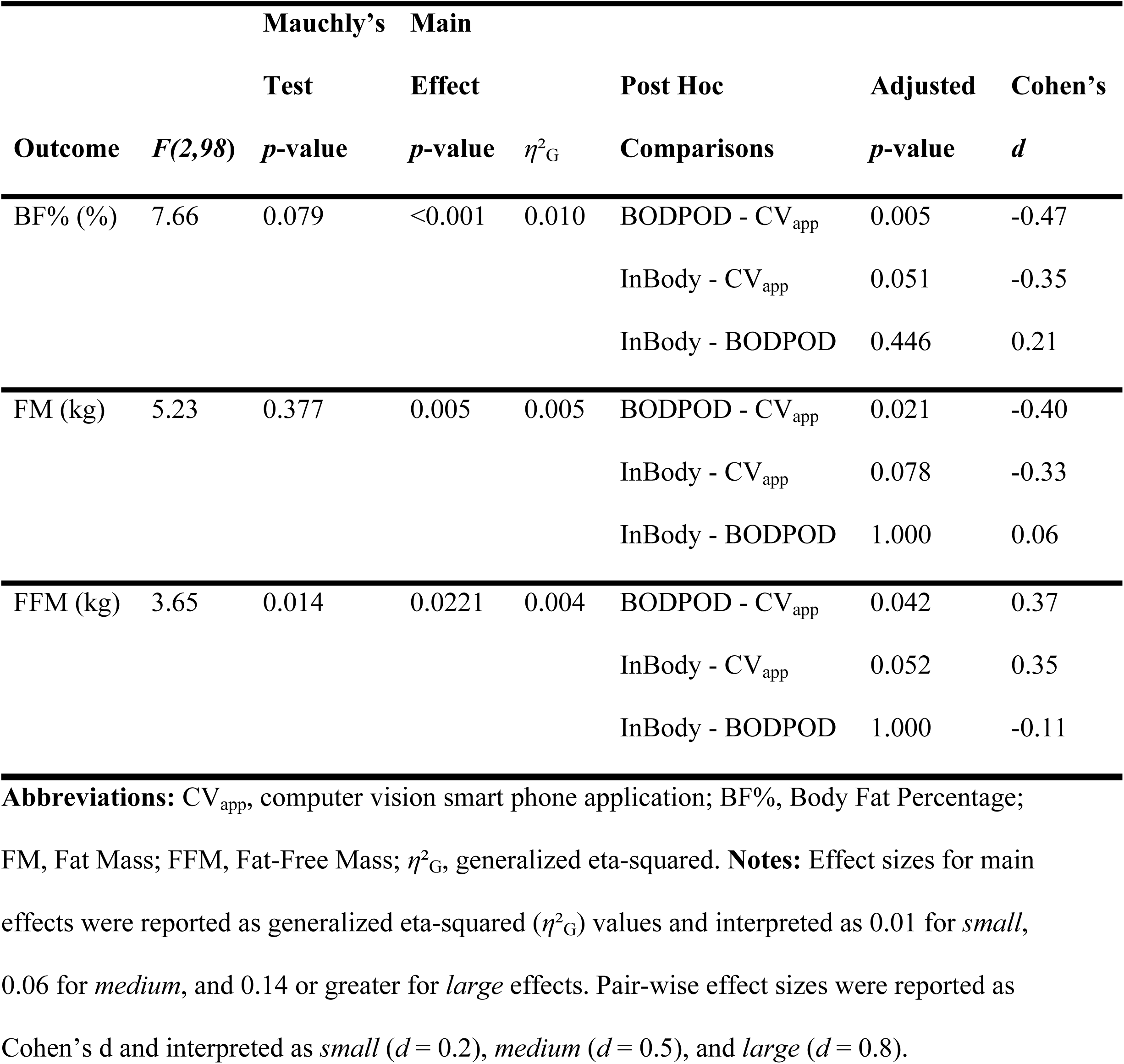
Repeated Measures ANOVA Results and Post Hoc Comparisons for Body Composition Measures.

Bland-Altman analyses supported differences between methods for BF%, FM, and FFM (Fig 2). Notably, wider LOA were observed when comparing the CV_app_ to BODPOD and InBody measures. For BF%, the CV_app_ overestimated BODPOD and InBody values (BODPPOD: mean bias = -2.2%, lower LOA = -11.2%, upper LOA = 6.8%; InBody: mean bias = -1.4%, lower LOA = -9.5%, upper LOA = 6.6%)with no evidence of proportional bias (*p* > 0.50 in both cases). For FM, the CV_app_ showed negative biases with wide LOA compared to both BODPOD (mean bias = −1.5 kg, lower LOA = -8.3 kg, upper LOA = 5.5 kg) and the InBody (mean bias = −1.2 kg, lower LOA = -8.6 kg, upper LOA = 6.1 kg), reflecting greater slightly larger measurement variability. No proportional bias was detected for these comparisons (*p* > 0.20). For FFM, the CV_app_ overestimated values relative to BODPOD (mean bias = 1.3 kg, lower LOA = -5.7 kg, upper LOA = 8.4 kg) and InBody (mean bias = 1.8 kg, lower LOA = -8.4kg, upper LOA = 12.0 kg). The BODPOD versus CV_app_ comparison indicated statistically significant proportional bias (*p* = 0.013), suggesting systematic error depending on FFM magnitude.

**Fig 2.**
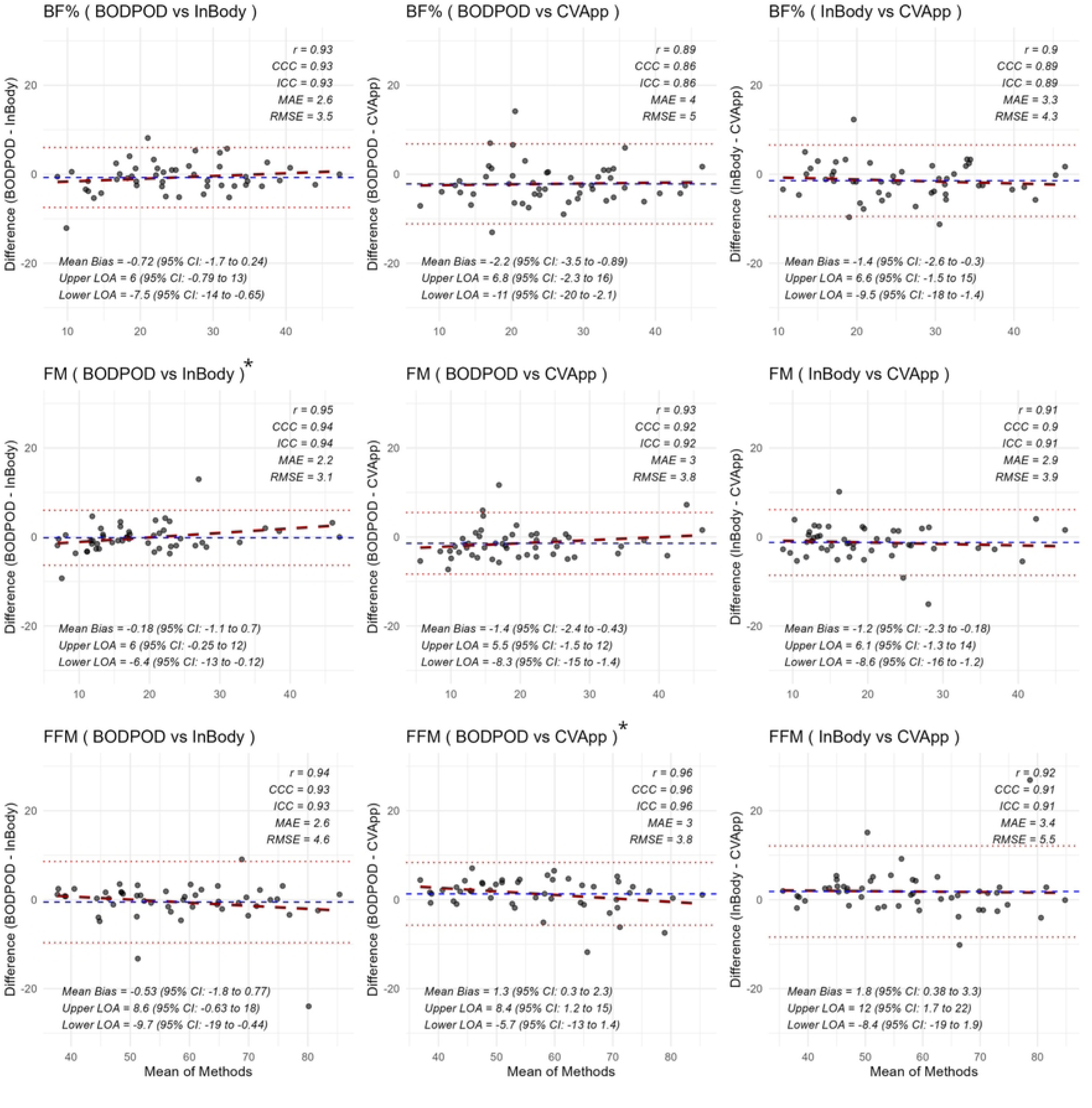
Bland Altman Plots Comparing Body Composition Methods **Notes:** Dashed blue line denotes the mean bias. The dotted red lines denote the upper and lower limits of agreements. The dashed red line indicates the regression of the difference in methods versus the body composition metrics, which indicates the proportional bias. Asterisk at subplot title denotes statistically significant proportional bias (*p* < 0.05). Mean bias, limits of agreement (LOA), and 95% confidence intervals (CI) are presented in the bottom left of each plot. Additional agreement statistics include Pearson’s r (*r*), Concordance Correlation Coefficient (CCC), intraclass correlation coefficient (ICC), mean absolute error (MAE), and root mean square error (RMSE). Thresholds for interpreting agreement statistics are as follows: 1) CCC: values < 0.90 = *Poor*, 0.90–0.95 = *Moderate*, > 0.95 = *Excellent* agreement; 2) ICC (2,*k*): values < 0.50 = *Poor*, 0.50–0.74 = *Moderate*, 0.75–0.89 = *Good*, ≥ 0.90 = *Excellent* reliability. **Abbreviations:** CV_app_, computer vision smart phone application; CCC, Concordance Correlation Coefficient; ICC, Intraclass Correlation Coefficient; MAE, Mean Absolute Error; RMSE, Root Mean Squared Error; CV_app_, computer vision smart phone application; BF%, Body Fat Percentage; FM, Fat Mass; FFM, Fat-Free Mass.

Additionally, agreement metrics comparing body composition methods are presented on Fig 1. Agreement metrics indicated that the CV_app_ had lower concordance with BODPOD compared to InBody vs BODPOD across all body composition measures. While ICC values were generally high (≥0.86) across all comparisons the CCC indicated poor agreement between the CVapp with both the BODPOD and InBody. Additionally, error metrics (MAE, RMSE) were relatively similar when comparing CV_app_ to BODPOD versus InBody to BODPOD.

### Test-Retest Reliability

Test–retest reliability metrics for each method are presented in Table 3. All methods demonstrated excellent reliability across all outcome measures, with ICCs exceeding 0.99. The CV_app_ exhibited the highest ICC values across the three methods, indicating near-perfect agreement between repeated measurements and also demonstrated the lowest SEM and CoV. The InBody SEM and CoV values were slightly higher than those of the CV_app_ but consistently lower than the BODPOD for all three outcomes. The BODPOD, while still demonstrating excellent reliability, had comparatively higher variability to the other methods. Interestingly, the BODPOD had the highest CoV observed for BF% (2.7%) among all methods.

**Table 3.** Test-Retest Reliability of Methods.

| Method | Metric | ICC | SEM | CoV | CCC | CR |
| --- | --- | --- | --- | --- | --- | --- |
| BODPOD | BF% | 0.995 | 0.7% | 2.7 | 0.995 | 0.02 |
|  | FM | 0.998 | 0.5 | 2.5 | 0.997 | 1.39 |
|  | FFM | 0.998 | 0.5 | 0.8 | 0.998 | 10.07 |
| InBody | BF% | 0.999 | 0.3% | 1.1 | 0.999 | 0.01 |
|  | FM | 0.999 | 0.3 | 1.1 | 0.999 | 0.70 |
|  | FFM | 1.00 | 0.3 | 0.4 | 1.000 | 0.70 |
| CV<sub>app</sub> | BF% | 1.00 | 0.2% | 0.6 | 1.000 | 0.00 |
|  | FM | 1.00 | 0.1 | 0.6 | 1.000 | 0.37 |
|  | FFM | 1.00 | 0.1 | 0.2 | 1.000 | 0.37 |

**Abbreviations:** CV<sub>app</sub>, computer vision smart phone application; BF%, Body Fat Percentage; FM, Fat Mass; FFM, Fat-Free Mass; ICC, Intraclass Correlation Coefficient; SEM, Standard Error of Measurement; CoV, Coefficient of Variation (%); CCC, Concordance Correlation Coefficient; CR, Coefficient of Reliability.

### Effects of Sex on Agreement Between Methods

The aggregate body composition data by sex and method are visually presented in Fig 3.

**Fig 3.**
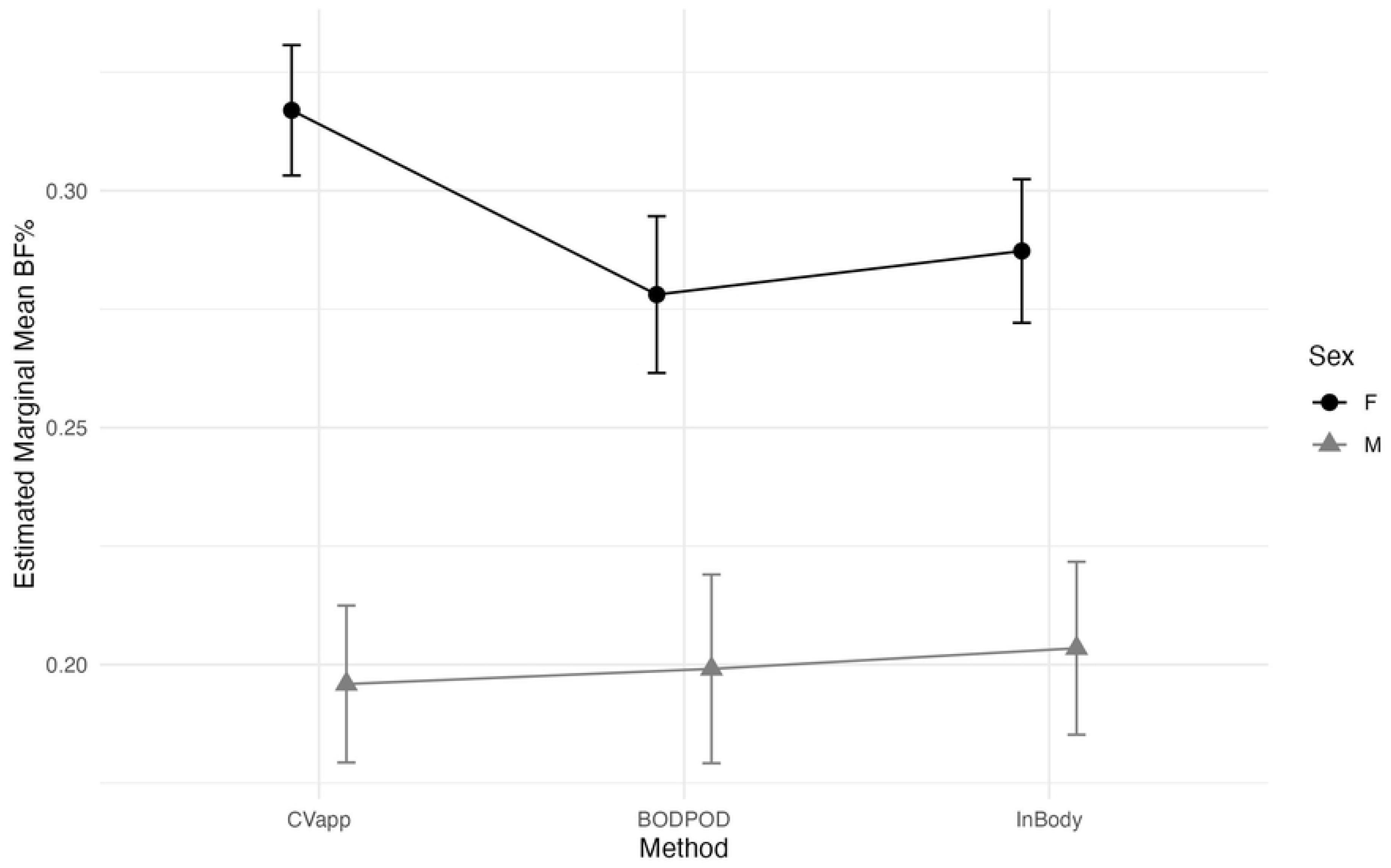
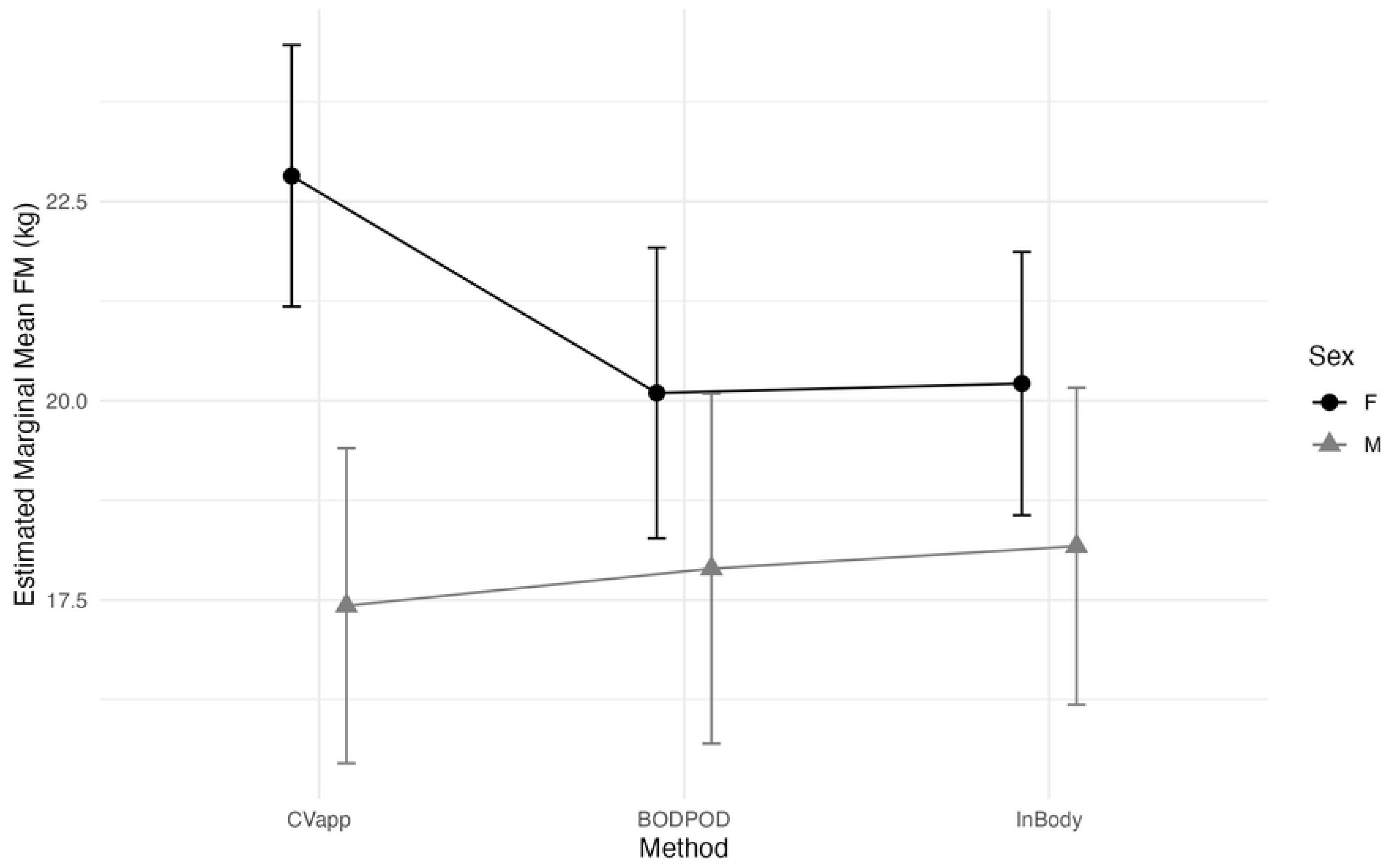
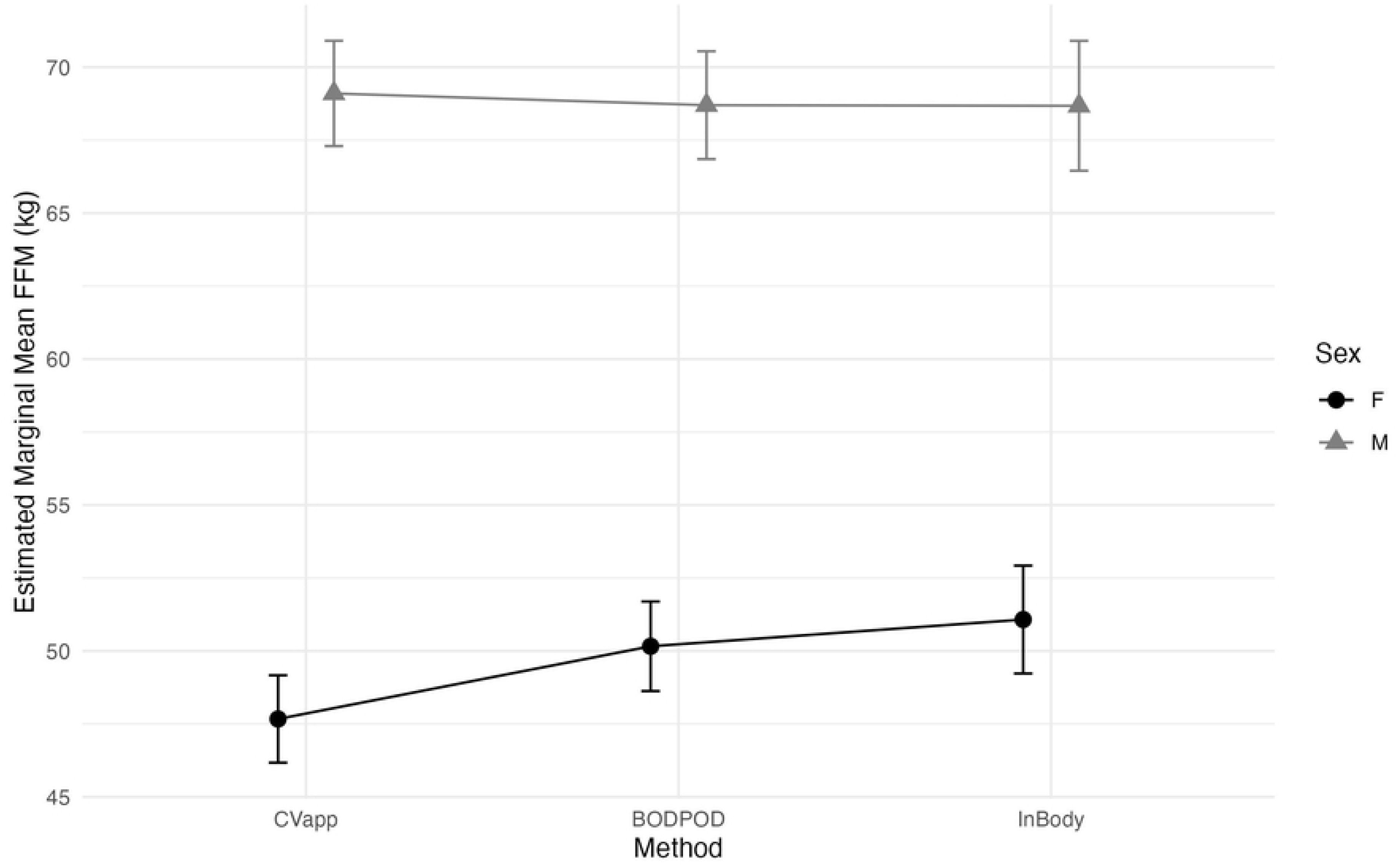
Interaction of Method and Sex on Body Composition Metrics A) Body Fat Percentage B) Fat Mass C) Fat-free Mass **Abbreviations:** CV_app_, computer vision smart phone application; M, Male; F, Female; BF%, body fat percentage; FM, fat mass; FFM, fat-free mass.

#### Body Fat Percentage

Significant main effects were found for Sex (*F*(1,47) = 17.04, *p* < 0.001, *η*²_G_ = 0.252) and Method (*F*(1.92, 90.22) = 5.40, *p* = 0.007, *η*²_G_ = 0.008) on BF%, as well as a significant Sex × Method interaction (*F*(1.92, 90.22) = 8.82, *p* < 0.001, *η*²_G_ = 0.013). Post-hoc comparisons showed that among females, the CV_app_ produced higher BF% values than both the BODPOD (mean difference = 3.9%, *p* < 0.001) and InBody (mean difference = 3.0%, p < 0.001), whereas there no differences existed between BODPOD and InBody (mean difference = 0.9%, *p* = 0.340). In males, no differences in BF% were observed between methods (all *p* > 0.630).

#### Fat Mass

Significant main effects were found for method on FM (*F*(1.98, 93.16) = 3.22, *p* = 0.045, *η*²_G_ = 0.003). There was no significant main effect of Sex, (*F*(1, 47) = 1.51, *p* = 0.226, *η*²_G_ = 0.030). However, the interaction between Sex × Method was significant (*F*(1.98, 93.16) = 7.94, *p* < 0.001, *η*²_G_ = 0.007), indicating that the difference in FM estimates across methods varied by sex. Post hoc comparisons within sex showed that, in females, the CV_app_ overestimated FM compared to both the BODPOD (mean difference = 2.72 kg, *p* < 0.001) and InBody (mean difference = 2.60 kg, *p* < 0.001), while BODPOD and InBody did not significantly differ. In males, no significant differences were found between any of the methods (*p* > 0.59). Comparisons between sexes within each method indicated a significant sex difference only for the CV_app_, with females having higher FM estimates than males (mean difference = 5.39 kg, *p* = 0.041). No significant sex differences were observed for BODPOD or InBody estimates.

#### Fat-free Mass

A significant main effect of Sex was found for FFM (*F*(1, 47) = 61.31, *p* < 0.001, *η*²_G_ = 0.545), with males having greater FFM than females. The main effect of Method only approached significance (*F*(1.69, 79.62) = 2.90, *p* = 0.069, *η*²_G_ = 0.005). However, a significant interaction between Sex × Method was observed (*F*(1.69, 79.62) = 4.94, *p* = 0.013, *η*²_G_ = 0.009), indicating that the difference in FFM estimates between methods varied by sex. Post hoc comparisons within sex indicated that among females, the CV_app_ underestimated FFM compared to both the BODPOD (mean difference = −2.49 kg, *p* < 0.001) and InBody (mean difference = −3.41 kg, *p* = 0.002), while BODPOD and InBody did not differ. Among males, no differences were found between any of the methods (*p* > 0.85). Across all three methods, females had lower FFM than males, with sex differences ranging from −17.6 kg to −21.4 kg (all *p* < 0.001), consistent with physiological expectations.

### Effects of Race and Ethnicity on Agreement Between Methods

The aggregate body composition data by race and ethnicity and method are visually presented in Fig 4.

**Fig 4.**
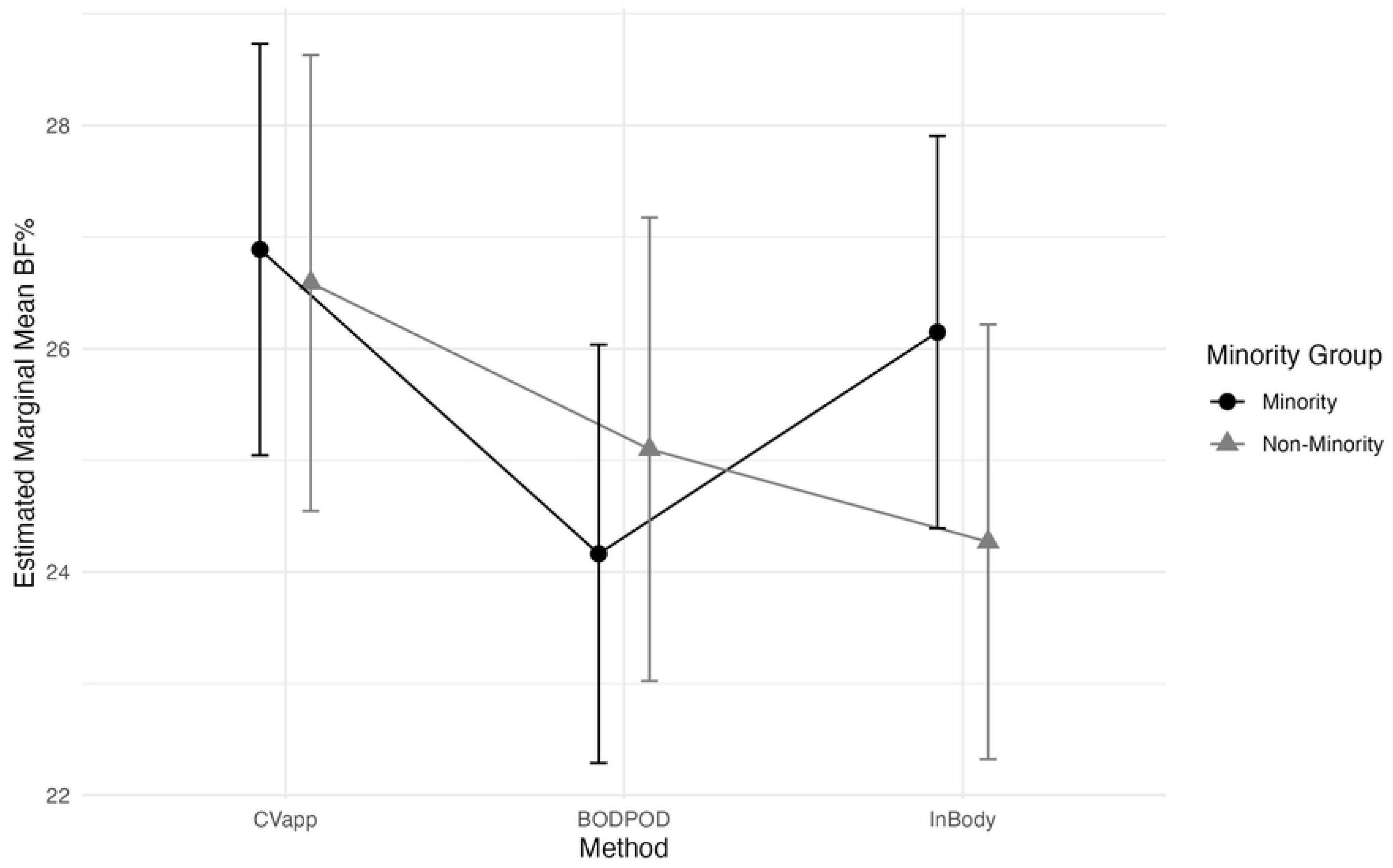
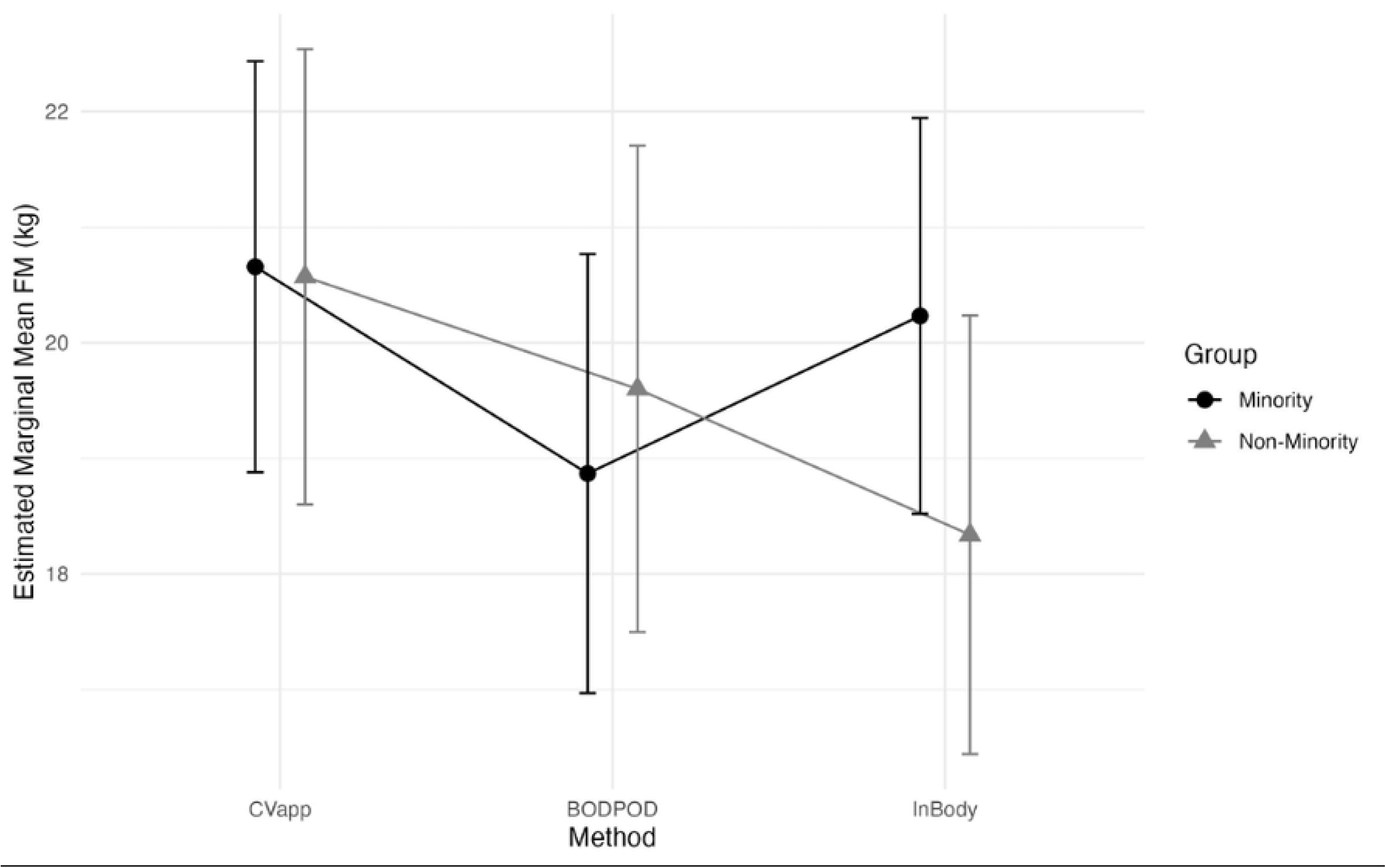
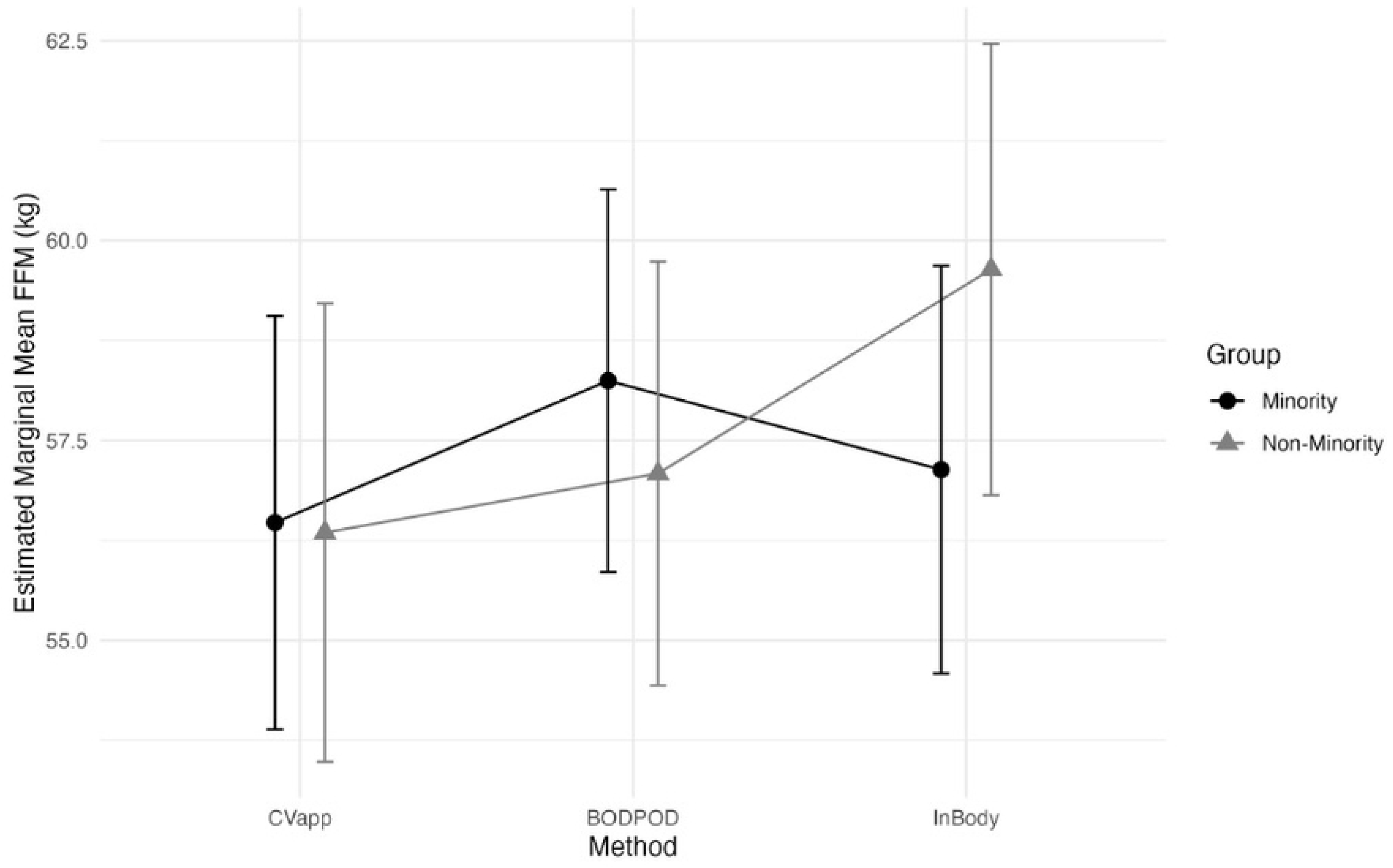
Interaction of Method and Racial and Minority Group on Body Composition Metrics A) Body Fat Percentage B) Fat Mass C) Fat-free Mass **Abbreviations:** CV_app_, computer vision smart phone application; BF%, body fat percentage; FM, fat mass; FFM, fat-free mass.

#### Body Fat Percentage

There was a significant main effect of Method (*F*(2, 94) = 7.25, *p* = 0.001, *η*²_G_ = 0.009), with BF% estimates differing across methods. The main effect of Minority Group was not significant (*F*(1, 47) = 0.02, *p* = 0.88, *η*²_G_ < 0.001), and although the Method × Minority Group interaction was not significant (*F*(2, 94) = 3.04, *p* = 0.053, *η*²_G_ = 0.004), there was a trend toward differences in measurement bias by Minority Group. Post hoc comparisons revealed that among Minority participants, BF% values from the CV_app_ were higher than BODPOD (+2.73%, *p* = 0.009), and BODPOD was lower than InBody (–1.99%, *p* = 0.006). Among Non-Minority participants, BF% from the CV_app_ was higher than InBody (+2.32%, *p* = 0.027). However, BF% did not differ significantly between groups within any individual method (all *p* = 0.47).

#### Fat Mass

There was no significant main effect of Minority Group on FM (*F*(1, 47) = 0.03, *p* = 0.875, *η*²_G_ < 0.001), indicating that average FM did not differ between groups. However, a significant main effect of Method was observed (*F*(2, 94) = 5.15, *p* = 0.008, *η*²_G_ = 0.005) and this was qualified by a significant Minority Group × Method interaction (*F*(2, 94) = 3.80, *p* = 0.026, *η*²_G_ = 0.004), suggesting that the differences among methods varied by group. Post-hoc pairwise comparisons revealed that, among Minority participants, FM estimates from the CV_app_ were higher than BODPOD (mean difference = 1.79 kg, *p* = 0.031), and BODPOD was lower than InBody (mean difference = −1.36 kg, *p* = 0.047). There was no significant difference between CV_app_ and InBody (*p* = 0.822). Among Non-Minority participants, FM from CV_app_ was higher than InBody (mean difference = 2.23 kg, *p* = 0.018). However, comparisons between CV_app_ and BODPOD (*p* = 0.410) and BODPOD and InBody (*p* = 0.113) were not statistically significant. There were no significant differences in FM between Minority and Non-Minority participants within any method (all *p* > 0.46).

#### Fat-free Mass

There was no main effect of Minority Group (*F*(1, 47) = 0.01, *p* = 0.9, *η*²_G_ < 0.001), indicating that overall FFM did not differ by group. A significant main effect of Method was observed (*F*(2, 94) = 5.04, *p* = 0.008, *η*²_G_ = 0.004), and a significant Minority Group × Method interaction emerged (*F*(2, 94) = 4.49, *p* = 0.014, *η*²_G_ = 0.004), suggesting the differences among methods varied across groups. Among Minority participants, FFM was lower when estimated by the CV_app_ compared to BODPOD (mean difference = −1.78 kg, *p* = 0.035). No other pairwise comparisons were significant (*p* > 0.38). Among Non-Minority participants, FFM estimates were lower using the CV_app_ compared to InBody (mean difference = −3.29 kg, *p* = 0.011), and BODPOD also reported lower FFM than InBody (mean difference = −2.55 kg, *p* = 0.022). No difference existed between CV_app_ and BODPOD t (*p* = 0.602). No significant differences existed in FFM between Minority and Non-Minority groups within any of the three methods (all *p* > 0.51).

To assess whether sex differences could be influencing the significant effects observed due to Minority group a chi-square test was conducted. There was no significant association between sex and minority group status, (χ²(1, N = 49) = 0.00, *p* = 1.00). Thus, the distribution of participants across sex (F, M) was similar for Minority (16 F, 11 M) and Non-Minority (13 F, 9 M) groups.

## Discussion

The primary purpose of this study was to evaluate the agreement and reliability of a smartphone-based CV_app_ for estimating body composition by comparing it to commonly utilized methods and to assess whether average values and r agreement differed by sex and racial/ethnic group. For the overall sample, the CV_app_ overestimated BF% and FM while underestimating FFM compared to both the BODPOD and InBody. However, subgroup analyses revealed that these differences were influence by sex and minority group status. Significant differences in body composition metrics were found for females but not males. The interaction between Minority Group and Method indicated that CV_app_ estimates were less accurate for Minority participants. Specifically, CV_app_ overestimated FM and underestimated FFM compared to BODPOD among Minority individuals, but not among Non-Minority participants. Interestingly, the interaction of Minority Group and Method did not reach statistical significance for BF%. The CV_app_ was found to have excellent test–retest reliability, indicating that the CV_app_ produces highly repeatable measurements even though those measurements have larger error between the other two measures in certain subgroups.

Previous studies of smartphone computer-vision body composition tools have generally reported high accuracy and minimal bias [20,21,23,24,26]. Notably, Majmudar et al. [26] evaluated a 2D (2 Dimensional) digital image smartphone application versus DXA in 134 adults and found very low MAE in BF% (∼2–3%) across the entire sample. Bland–Altman analysis showed computer vision had no significant bias and the narrowest limits of agreement (LOA; ±5% fat) with DXA, in contrast to the other methods which exhibited systematic biases and wider variability [26]. Errors were only slightly higher in females (MAE = 2.3%) than men (MAE = 1.9%), and modestly higher in Black versus White participants (BF% MAE 2.7% vs 2.0%) [26]. In that study, the VBC app showed strong concordance with DXA for both sexes and no significant systematic error by demographic subgroups [26]. As previously described, Farina et al. [24] and Affuso et al. [23] also reported evidence supporting that 2D digital image approaches obtained with a smartphone camera can be used to estimate BF%, with strong correlation to DXA BF% values. Similarly, A 3D smartphone-scanning approach (non-rigid avatar reconstruction) also yielded extremely high reliability (ICC=0.996–0.997) and showed no evidence of proportional bias in BF% estimates for two BF% estimation equations [27]. Tinsley and colleagues found that two estimation equations were statistically equivalent to DXA within a ±2% margin and showed strong linear associations (Pearson’s r and CCC = ∼0.90) [27]. The average MAE was approximately 3.5%, with a SEM around 4.2% across equations [27].

In contrast, Graybeal et al. [18] evaluated the validity and reliability of body composition estimates generated by smartphone applications against a 4-compartment criterion model in a diverse sample of adults (n=184; 114F, 70M). In this study only one smartphone application (HALO) demonstrated equivalence for BF% with the 4-compartment model, while all smartphone applications showed proportional bias for BF% and varying degrees of error for FM and FFM [18]. Notably, the accuracy of body composition estimates from smartphone applications varied by sex and racial/ethnic group, likely due to differences in the underlying algorithms and the demographic composition of the data used to train them [18]. It should be noted that the smartphone CV_app_ used in the present study was not investigated in the aforementioned studies, which suggests that machine learning algorithm and training data could explain differences in the findings. Despite these limitations in validity, all smartphone application based methods demonstrated excellent reliability (ICCs ≥ 0.99) [18]. Together, the prior findings [18,26,27] when combined with the current study findings suggest that while smartphone CV_app_’s to assess body composition can be highly reliable and accurate in ideal conditions, the results can vary depending on the application used and the user demographic characteristics.

### Practical Implications

The body composition estimation biases we noted in the current study have important practical implications. In consumer health or clinical settings, biases could result in inappropriate advice or loss of trust in the technology. The CV_app_’s high reliability means it could be useful for tracking relative changes over time (e.g. weight-loss progress) within an individual. However, because the absolute values were biased, practitioners should be cautious using this CV_app_ for baseline assessments or cross-sectional comparisons. Specifically, overestimating BF% in females and minorities could lead to misclassification of health risk or unnecessary concern, while underestimating FFM might obscure loss of lean mass. Furthermore, the demographic bias could raise equity concerns. AI-driven tools that underperform for certain groups can exacerbate health disparities [33,34]. For example, facial-recognition studies have shown substantially smaller error rates for light-skinned males versus dark-skinned females [35,36]. In the context of the current study, a body-composition app that is less accurate for females and minorities risks may perpetuate unequal care. Health technology developers and regulators should thus require subgroup validation to ensure objectivity before wide deployment [33,34]. Despite these issues, computer vision-based methods offer clear advantages (e.g., low cost, accessibility, no radiation, etc.) and could serve as screening or monitoring tools when more advanced methods (e.g., DXA, MRI) are unavailable [37]. For example, digital anthropometry has been advocated as a convenient pre-screening step [37]. Our results suggest, however, that any such screening should come with caveats, which clinicians and users must be aware that certain groups may get systematically skewed results.

### Limitations and Future Research Directions

This study has certain limitations that warrant consideration. While both the BODPOD and InBody are frequently used in research studies, they have known errors and limitations [13,16,38]. For instance, the BODPOD can overestimate BF% in very lean individuals and underestimate BF% in obese subjects [39]. Thus, some of the CV_app_’s discrepancy might partly reflect reference-method limitations. Future work could compare the CV_app_ directly to DXA [26,27] or 4C [18] measurements to isolate algorithm error. We also note that our sample size could be considered limited [18,26,27], which could affect generalizability. The observed subgroup biases likely stem from the CV_app_’s algorithm and training data. If the underlying neural network was trained on a dataset skewed toward lighter-skinned or male bodies, it may not generalize well to females or minorities. In our case, variations in skin tone could also affect how the computer vision segments the body from background. Darker skin has lower contrast against certain backgrounds, and cameras have nonlinear color response, which can introduce systematic segmentation errors. Another limitation of this study was the dichotomous classification of participants as either racial/ethnic minorities or non-minorities. Because skin pigmentation can vary widely within these broad categories, future research should measure and analyze actual skin pigmentation rather than relying solely on racial or ethnic identification. Females and different ethnic groups can have distinct fat distributions [40,41]. For example, woman may have more subcutaneous fat in hips/thighs for females and visceral vs subcutaneous ratios can vary by ancestry [40,41]. A 2D silhouette-based algorithm might not capture these patterns equally for all body types. If the CV_app_’s model assumes a certain body shape, it could incorrectly estimate in bodies that deviate from that norm. Further development should include diversifying the training set to include more females and minority body images under varied lighting/clothing conditions. Algorithmic fixes might involve multi-spectral imaging or depth sensing to reduce reliance on visible-light contrast [42]. Rigorous diversity and equity audits (e.g., testing on curated groups) should be standard practice during CV_app_ validation processes. Furthermore, expanding validation to larger, more varied cohorts, including extremes of BMI, older adults, different clothing, will help characterize and correct biases.

## Conclusion

In summary, the current study highlights both the promise and limitations of smartphone computer vision-based body composition technology. While the CV_app_ demonstrated excellent reliability, it showed significant demographic bias by overestimating BF% and FM, and underestimating FFM in female users. Study findings contrast with several prior studies [20,21,23,24,26] reporting unbiased performance and indicate that not all CV_app_ are equally robust across demographic profiles. Importantly, a CV_app_ can be reliable yet systematically inaccurate for specific populations. Health professionals and researchers should interpret CV_app_ estimates with caution, particularly when assessing diverse groups. Until algorithms are refined to eliminate bias, these tools should not serve as standalone diagnostic measures. Future research should prioritize inclusive datasets and algorithm transparency to ensure equitable performance as digital anthropometry continues to evolve. These steps will be critical to ensure emerging digital health tools contribute positively to health equity, rather than inadvertently widening disparities.

## Data Availability

All relevant data are within the manuscript and its Supporting Information files. The dataset generated and analyzed during the current study has been deposited in the Open Science Framework and is publicly available under DOI 10.17605/OSF.IO/RUZ2W (version 1.0). The dataset can be accessed at: https://osf.io/ruz2w/?view_only=5b33e65b7a3c4e33b4dd53f0371338aa

https://doi.org/10.17605/OSF.IO/RUZ2W

https://osf.io/ruz2w/?view_only=5b33e65b7a3c4e33b4dd53f0371338aa

## Acknowledgements

None.

## Notes

### Competing Interest Statement

The authors have declared no competing interest.

### Funding Statement

The author(s) received no specific funding for this work.

### Author Declarations

The study procedures were approved by George Mason University (IRB #: 1665548).

